# An ML prediction model based on clinical parameters and automated CT scan features for COVID-19 patients

**DOI:** 10.1101/2022.01.30.22269998

**Authors:** Abhishar Sinha, Swati Purohit Joshi, Purnendu Sekhar Das, Soumya Jana, Rahuldeb Sarkar

## Abstract

Outcome prediction for individual patient groups is of paramount importance in terms of selection of appropriate therapeutic options, risk communication to patients and families, and allocating resource through optimum triage. This has become even more necessary in the context of the current COVID-19 pandemic. Widening the spectrum of predictor variables by including radiological parameters alongside the usually utilized demographic, clinical and biochemical ones can facilitate building a comprehensive prediction model. Automation has the potential to build such models with applications to time-critical environments so that a clinician will be able to utilize the model outcomes in real-time decision making at bedside. We show that amalgamation of computed tomogram (CT) data with clinical parameters (CP) in generating a Machine Learning model from 302 COVID-19 patients presenting to an acute care hospital in India could prognosticate the need for invasive mechanical ventilation. Models developed from CP alone, CP and radiologist derived CT severity score and CP with automated lesion-to-lung ratio had AUC of 0.87 (95% CI: 0.85-0.88), 0.89 (95% CI: 0.87-0.91), and 0.91 (95% CI: 0.89-0.93), respectively. We show that an operating point on the ROC can be chosen to aid clinicians in risk characterization according to the resource availability and ethical considerations. This approach can be deployed in more general settings, with appropriate calibrations, to predict outcomes of severe COVID-19 patients effectively.

## Introduction

The COVID-19 pandemic, caused by the coronavirus Sars-Cov-2, has been ravaging countries around the world since early 2020 in waves dominated by different variants. As recently as on 26th of November 2021, another newly declared variant of concern (VOC) called Omicron^1^ has caused significant alarm across the globe with multiple significant mutations, reminding us that the pandemic and its associated concerns are far from over. Around the peaks of each wave, healthcare systems were stretched, as large number of people simultaneously required urgent medical attention in the form hospitalization, and some of them required admission to the intensive care units for mechanical ventilation, which is extremely scarce resource^2,3^. In this backdrop, prioritization and triaging became a key area for discussion in order to ensure that the care is directed to patients who require it most and benefit the most from it. Multiple ethical guidelines have been published based on clinical parameters, including guidance for optimizing allocation of mechanical ventilators, even in developed economies^4,5^. However, in a high volume patient admission scenario, risk stratification for clinically meaningful outcomes of admission to hospital, intensive care units and/or mechanical ventilation may be challenging and potentially suboptimal, when the model is based on clinical parameters alone, as it misses out on vital data derived from radiological examination.

Machine learning (ML) models have been used in the development of prognostic models already in the current pandemic^6,7^. CT imaging, that can provide us with detailed involvement of lung tissue and the contiguous structures in the disease process with reasonable detail, can be added to the clinical parameter based models to make the prognostication more comprehensive. It may be noted that severity of lung infection has already been measured using the CT severity score based on chest CT scans, and this requires manual assessments by a radiologist^8^. This is vulnerable to intra- and inter-observer variability^9,10^, especially at a time of high volume reporting. In fact, a prediction model has been developed with amalgamation of clinical and radiological features^11^. However, the radiological quantification of disease was done manually in that model, which can be labor intensive and subject to the variabilities mentioned above. Additionally, in low and middle income countries (LMIC) such as India, qualified radiologists are scarce relative to the population. For example, India has a meagre 0.9 physicians per 1000 population^12^, of whom only a small proportion are radiologists, making consistently high quality assessment of radiological images a challenge. In such situations, an automated method to quantify the inflammation in the lungs can be useful to reduce the workload on the radiologists with potentially improved consistency in reporting. We hypothesize that first, algorithmic scoring of CT scans could be almost as effective as manually generated CT severity scores in delineating pulmonary disease activity, and second, this can in turn contribute towards a prognostication model in conjunction with clinical parameters. Therefore, demographic and clinical data such as age and sex, blood-based biomarkers such as C reactive protein (CRP) and D dimer, and pre-existing conditions such as diabetes and hypertension, which have all been shown to prognosticate COVID-19, can be combined with an algorithmically generated lung inflammation index, in order to make a clinically robust prognostic model in predicting clinically meaningful outcomes, of which we considered the need for invasive mechanical ventilation as the outcome of our choice in view of it’s importance for the severely affected patients in the background of resource scarcity. An important point to consider would be the need for re-calibration of the model in different scenarios in order to avoid bias that may arise from differences in patient demographics and unique healthcare delivery in each setting. Currently, there is a lack of model development in LMICs, an issue which the current study can potentially address.

In the current study, we have addressed the two aforementioned important issues by developing a ML-based clinico-radiological outcome prediction model on patients based in a university hospital in western India. The measurement of the disease burden in the CT scan has been automated, which reduces the workload of reporting radiologists.

## Methods

The study was approved by the Institute Review Board of Mahatma Gandhi Medical College and Hospital (MGMCH), Jaipur, India (Contact Institutional Ethics Committee MGMCH). The informed consent was waived due to the retrospective observational characteristic of the study by the Review Board. All methods were performed in accordance with the relevant guidelines and regulations of the Review Board of MGMCH.

We developed an automated system to predict if a patient with COVID-19 is going to need mechanical ventilation, combining data from chest CT scan and clinical parameters. We algorithmically computed Automated Lesion Lung Ratio (ALLR) from the CT scan of each subject, and trained an ML prediction model using clinical features and ALLR. We also trained two other models using only clinical features and clinical features along with CT severity score, to compare the performance with the fully automated system, and verify that we are not losing accuracy while reducing the involvement of radiologists. A flow diagram to show the method is shown in Figure 1.

**Figure 1.**
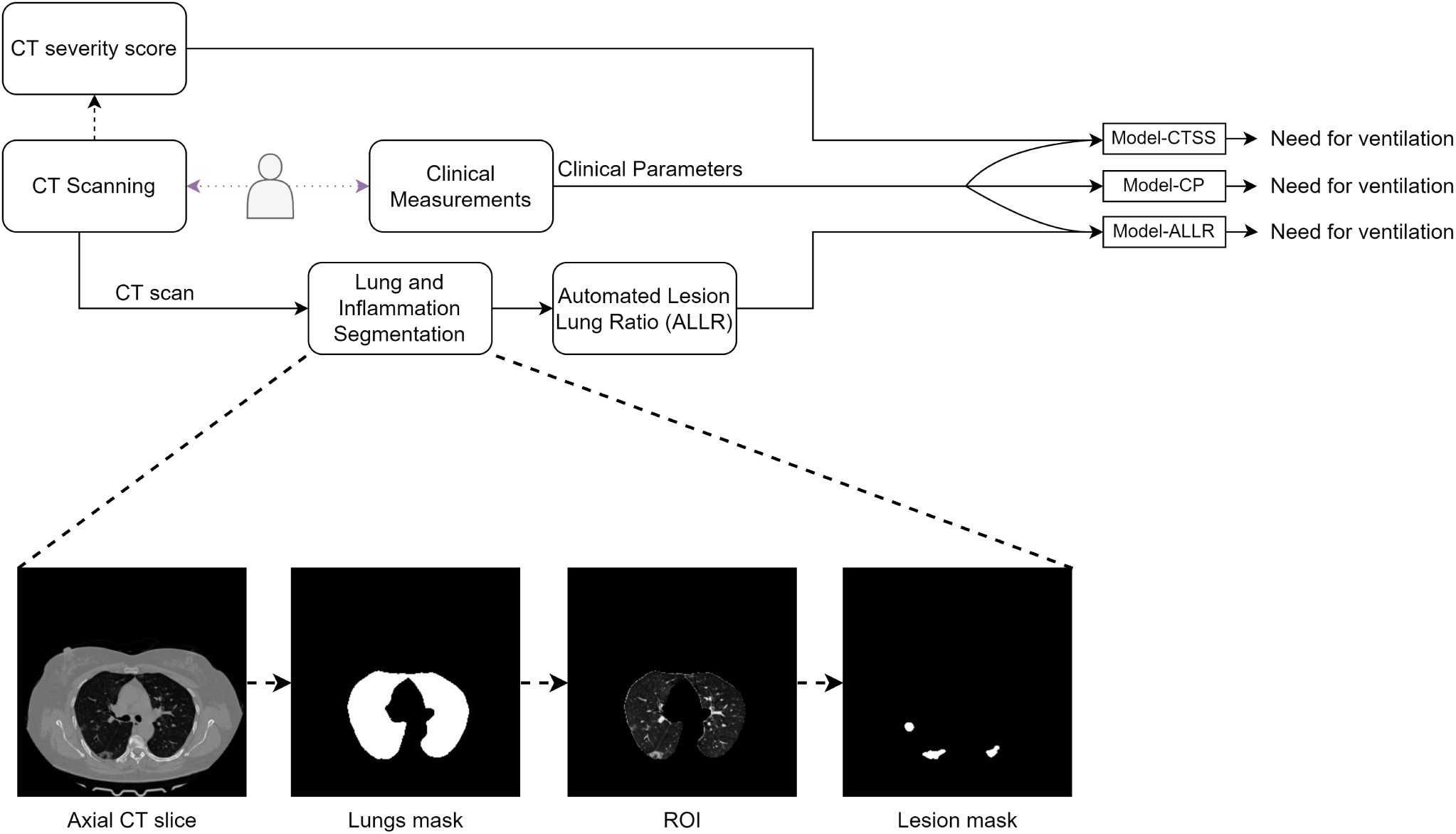
Flowchart. Top panel shows the flow chart of the project. This shows the experiment of training 3 models on different feature sets. Model-CP was trained only on clinical parameters. Model-CTSS was trained on clinical features appended with CT severity score. Model-ALLR was trained on clinical features appended with ALLR. The bottom panel is the expanded view of lung and inflammation segmentation. The lungs mask are produced by a segmentation model from CT scan. This mask is used to get the region of interest (ROI). The ROI is given as input to another segmentation model that produces the inflammation masks.

Here, to compute ALLR, we needed to segment the lungs and their inflamed regions. For such segmentation tasks, we needed to train two different ML models based on separate annotated datasets.

### Dataset Description

To train the segmentation models, we relied on publicly available datasets. For the lung segmentation task, we made use of a subset^13^ of the COVID-19 CT Lung and Infection Segmentation Dataset^14^. This dataset, henceforth called dataset-1, has 2581 axial slices of CT scans from 10 subjects, each consisting of non-contrast CT scans with a slice thickness of 1-1.2mm and slice distance of 1-1.2mm, as well as the corresponding lung masks. For the inflammation segmentation task, we used the dataset (henceforth referred as dataset-2) from COVID-19 Lung CT Lesion Segmentation Challenge^15–17^, consisting of 13705 axial slices of chest CT scans from 199 patients taken at slice thickness 5mm and slice distance 5mm, and the corresponding inflammation masks. The data preparation details are given in Supplementary section 1.

Further, clinical and radiological data of COVID-19 patients were collected at MGMCH between October to December 2020 for the purpose of developing and validating the prognosis model. The data included anonymized CT scans as well as clinical, demographic, biochemical, and radiological parameters of 302 COVID-19 patients. Volumetric thin section CT scans were obtained with slices of 0.625mm each with high spatial frequency. The clinical parameters in the dataset and other details are furnished in Table 1. This dataset also had missing values, these were replenished via data imputation. Specifically, the missing day of presentation was imputed with the median value. Missing values for the rest of the features were imputed using the nearest neighbors method with the assumption that similar known parameters indicate similarity in unknown parameters as well^18^.

**Table 1.** Summary of parameters in the MGMCH dataset and the number of records that have the parameter recorded. ^a^ Median (interquartile range) ^b^ Number of days passed from the symptom onset

| Parameter | Number of records with parameter value included | Summary |
| --- | --- | --- |
| Age | 302 | 59 (40-79) <sup>a</sup> |
| Sex | 302 | 75.16% male |
| Day of presentation<sup>b</sup> | 264 | 4 (3.75-5) <sup>a</sup> |
| CT severity score | 302 | 15 (10-20) <sup>a</sup> |
| CRP | 202 | 34.9 (13.77-78.5) <sup>a</sup> |
| D Dimer | 191 | 278 (221.5-483) <sup>a</sup> |
| Ferritin | 159 | 343 (155.75-713.95) <sup>a</sup> |
|  |  | Prevalence |
| Diabetes | 300 | 19.53% |
| Obesity | 300 | 16.22% |
| Hypertension | 300 | 30.13% |
| Need for oxygen | 302 | 39.74% |
| ICU admission | 302 | 21.52% |
| Need for ventilation | 302 | 11.59% required |
| Death | 302 | 5.96% mortality |

### Model Development

We developed ML models to tackle two major tasks, namely, (i) lung and inflammation segmentation in CT images, and (ii) multiparameter prediction of the outcome.

#### CT Image Segmentation

In CT images, we first delineated the lungs, and subsequently marked the inflamed regions within the lungs. For each subtask, we make use of a suitable deep network.

##### Lung segmentation

For lung segmentation, we used Bi-directional Conv-LSTM U-Net with Densely Connected Convolutions (BCDU-Net), which had reported high performance in an analogous task in a lung cancer study^19^. Inspired by that work, we adopted a complementary approach, where, instead of a mask for the lungs, one uses that for the chest region excluding lungs. Further, we made certain improvements in the preprocessing step for generating such masks, by identifying areas for improvements by manual assessment. Specifically, trachea had wrongly been segmented as part of the lung. To prevent this without compromising the segmentation accuracy, we performed a convex hull operation that caused the trachea to now be correctly identified. Training and test subsets were generated by randomly splitting dataset-1 (10 CT scans) in a 4:1 ratio. We made use of the intersection over union (IoU) loss criterion (i.e., 1 - IoU), and the Adam optimizer with an initial learning rate of 10^*−*4^. The model was trained for 100 epochs. In view of the small size of the dataset, image augmentation was used while training as an aid to the generalizability of the model. Data augmentation details are presented in Supplementary section 2.1.

To improve the quality of lung masks generated by model, the following post processing steps were included in the lung segmentation algorithm. Morphological opening operation with a disk of size 3 pixels as structuring element was used to disconnect trachea from lung, and then connected components were labeled in the mask and the components with size less than a third of total volume were removed, so that only the lungs were present in the mask. We identified another qualitative issue with the model’s prediction, which was not evident from the dice score. The boundaries of the lungs were jagged because of misclassifications, in some cases where there was a thickening of the pleura or presence of peripheral ground glass. Morphological geodesic active contour^20,21^ was used to smooth the boundaries. The mask was given as an initial level set to the algorithm to minimize the internal energy function, which smoothens the boundaries of mask. The multipliers for balloon/pressure force and the internal energy terms were 1. The number of iterations was set to 10

##### Lung inflammation segmentation

For lung inflammation segmentation, we employed UNet++, which extends UNet by introducing a denser network of skip connections with convolution blocks bridging the semantic gap between the encoder and decoder feature maps^22^, and whose performance we compared with that of BCDU-Net. Training and test subsets were created by splitting dataset-2 in the ratio of 4:1. Noting that the gray-scale values vary widely within the manually annotated lesions (lighter shade indicating higher severity, while low-severity areas being close to uninfected tissues in shade), we sought to focus on subregions with severe infections in order to facilitate its automated distinction from healthy lung tissues. For this purpose, we initialized with the contour of original annotations, and shrank those by the morphological geodesic active contour method to high-severity subregions. The multipliers for the internal energy and pressure terms were 0 and -1.25, respectively. The number of iterations were set to 50.

We trained the inflammation segmentation model using such subregions as reference. As earlier, we minimized IoU loss using Adam optimizer with an initial learning rate of 10^*−*4^. The model was trained for 30 epochs, with data augmentation. The developed model was used to segment inflamed regions in each 2D axial slice of a CT scan. The data augmentation details are given in Supplementary section 2.2.

##### Performance index

We measured the segmentation fidelity in terms of Dice coefficient (DC) between the estimated and the reference regions, defined as twice the area of the intersection of the said regions divided by the sum of the areas of those regions. Stacking the aforementioned axial slices, we obtained the CT volume, in which the segmented lungs assembled into the lung volume, and the inflamed subregions into the lesion volume. The ALLR was subsequently computed as the volumetric ratio of lesion to lung. To each MGMCH data record, we appended the ALLR for subsequent analysis.

#### Outcome Prediction

While admitting each COVID-19 patient, the hospital records the demographic, the biochemical as well as the radiological parameters based on the chest CT scan (if performed) along with pre-existing conditions. In this study, we used the patient records to develop an ML model to predict the need for mechanical ventilation (MV), an acutely scarce resource, based on the MGMCH dataset. Prediction of such need is a clinically meaningful and significant outcome, which may guide efficient triaging (e.g., patients in need of MV should be admitted to a hospital in a unit where they can be monitored effectively).

##### ML model

Predicting the need for MV was posed as a classification task, and we assessed the efficacy of competing ML models for this task. Specifically, we compared the performance of random forest^23^ and XGBoost^24^ (extreme gradient boost), two well-known ensemble models, usually superior to single models in terms of generalizability and robustness^25,26^. The random forest, a bagging type of ensemble, consists of multiple independent decision trees, each of which is trained using a random subset of features and a random subset of samples drawn with replacement, and hence enjoys reduced variance of the ensemble, avoiding overfitting. In contrast, XGBoost makes use of a type of gradient boosting, where multiple decision tree models are trained in succession, each tending to improve performance. As a result, the ensemble, a weighted combination of component models, enjoys both reduced bias and reduced variance. We tuned the hyperparameters via grid search in case of both random forest and XGBoost.

In order to test if algorithmically generated ALLRs are as effective as the manually generated CT severity scores in predicting the need for ventilation, we performed the following experiment. We established as a baseline the performance of a reference ML model (via suitable training) based only on clinical parameters. Subsequently, two additional models were developed, where clinical parameters were appended with the CT severity score in case of one, and the ALLR in case of the other. Performance of those two models were compared under various criteria with the performance of the reference model as a baseline. Here, we considered two versions of each of the models, one based on random forest and the other based on XGBoost. For all models the categorical features were encoded using label encoding and the numeric features were treated as continuous variables.

In case of each model, we performed Monte-Carlo cross validation^27^.The dataset was randomly split into training and test subsets in a 4:1 ratio in a stratified manner. The model was trained on the former subset, and the outcome was predicted for each case in the unseen test subset. In particular, the probability of the outcome was recorded for each test case, which led to a binary decision based on a suitable threshold. To report various statistics involving test performance, this above process was repeated for 50 independently generated training-test partitions. Specifically, the model with the median performance may be taken as a representative.

##### Performance indices

The receiver operating characteristic (ROC) curve is plotted by calculating true positive rate (TPR) and false positive rate (FPR) for various threshold points of the probability generated by a model. ROC curve is convex by definition, so we took the convex hull of the curve, where the operating points between the optimal points are obtained by time sharing. We also calculate the area under the curve (AUC) to compare different models. It is a more useful performance metric than accuracy in cases of class imbalance, which is present in the MGMCH dataset.

Further, we computed the confusion matrix to compare different models. A confusion matrix includes TPR, FPR, true negative rate (TNR) and false negative rate (FNR) which are calculated after thresholding. Generally, the threshold probability is selected to maximize the accuracy. In different settings, TPR and TNR may not always have equal importance but accuracy weighs them equally. Hence, we also calculate confusion matrices using multiple thresholds selected by minimizing a weighted loss FPR + *α* FNR for different values of *α* We also visualize the different operating points on the ROC curve.

Facilitation of human decision making: In the above, the probability threshold *τ* may need to be adjusted to reflect the availability of various resources. However, the probability value predicted by the model may not easily be interpretable by clinicians. As a solution, we suggest a piecewise linear mapping from the machine-generated probability to an interpretable one as follows. Probabilities between 0 to *τ* were linearly transformed to probabilities 0-0.5, and probabilities from *τ* to 1.0 were mapped to probabilities 0.5-1.

#### Implementation Notes

Computer coding was carried out in Python programming language version 3.8^28^, and compute-resources from Google Colab^29^ were used. Deep learning models were implemented using TensorFlow v2^30^ library. OpenCV v4^31^ and scikit-image v0.17^32^ were used for image processing. We used random forest model from the scikit-learn v0.24^33^ library.

## Results

The demographic, physiological and outcome on mechanical ventilation data are presented in Table 1.

The lung segmentation had a Dice coefficient of 96% on the test set. Inflammation segmentation had a dice coefficient of 58% on the test set using BCDU-Net while UNet++ improved the Dice coefficient to 60%. Even though the Dice coefficient for inflammation segmentation is less compared to lung segmentation, in view of this being a somewhat subjective problem we proceeded to check if ALLR work as well as CT severity score.

The mean and standard deviation of AUCs calculated on the validation sets for the 50 iterations are given in Table 2. The random forest models exhibited higher mean AUCs than XGBoost models. The mean AUC for the random forest model with clinical features and ALLR was 0.91 which is greater than that of the random forest model using only clinical features. All the random forest models had a low standard deviation of 0.06 for AUC which indicated that the results are consistent. The radiological features were ranked as the most important feature for both models using ALLR and CT severity score as shown in Figure 2. The mean ROC curve for the 3 classification tasks using random forest models is given in Figure 3. It shows different operating points corresponding to minimizing different loss functions. The models including radiological features had higher TPR than the model using only clinical features at every operating point.

**Table 2.**
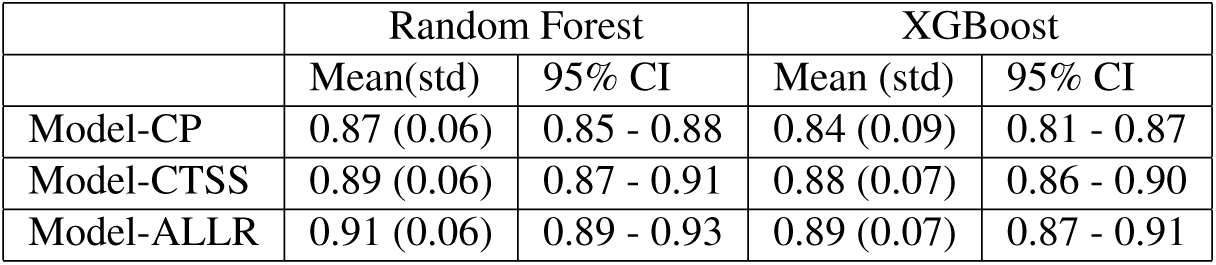
Validation AUCs for 50 iterations for the three models. Abbreviations: Model-CP: Model with clinical parameters as input, Model-CTSS: Model with clinical parameters and CT severity score as input, Model-ALLR: Model with clinical parameters and ALLR as input.

|  | Random Forest |  | XGBoost |  |
| --- | --- | --- | --- | --- |
|  | Mean(std) | 95% CI | Mean (std) | 95% CI |
| Model-CP | 0.87 (0.06) | 0.85 - 0.88 | 0.84 (0.09) | 0.81 - 0.87 |
| Model-CTSS | 0.89 (0.06) | 0.87 - 0.91 | 0.88 (0.07) | 0.86 - 0.90 |
| Model-ALLR | 0.91 (0.06) | 0.89 - 0.93 | 0.89 (0.07) | 0.87 - 0.91 |

**Figure 2.**
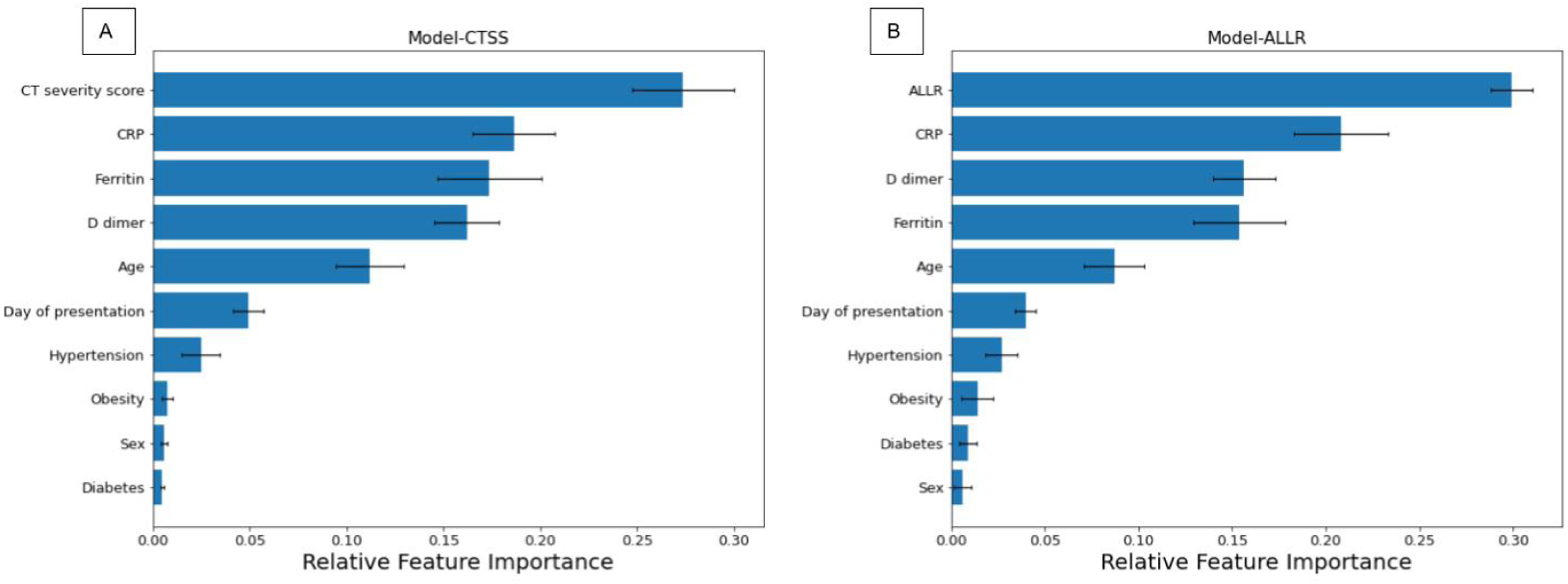
Relative feature importance. Panel A shows mean feature importance of the model having CT severity score and clinical parameters as the input. Panel B shows mean feature importance of the model having ALLR and clinical parameters as the input. The error bars show the standard deviations.

**Figure 3.**
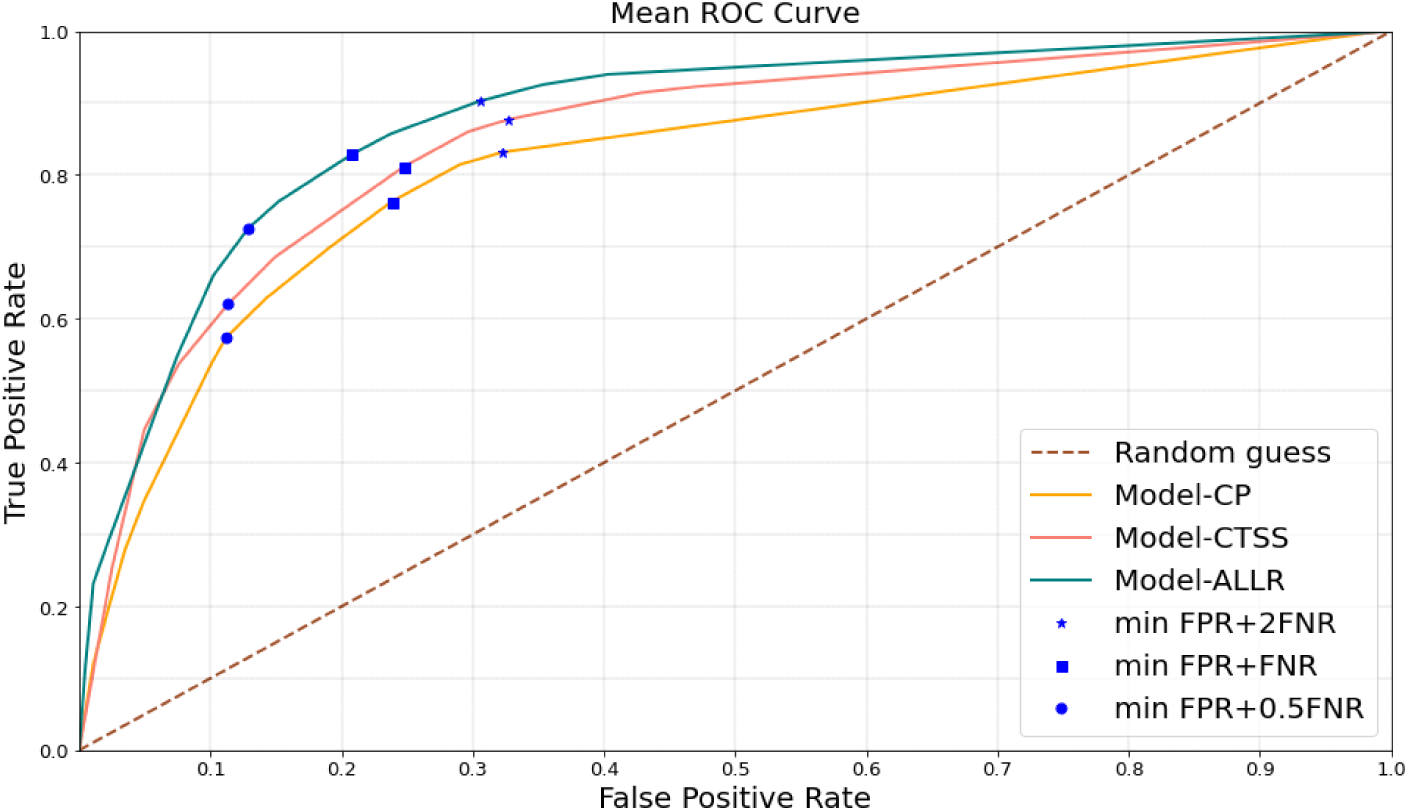
Mean ROC curve. The figure shows the ROC curves obtained by taking the mean of validation ROC curves over 50 iterations for predicting need for ventilation for the three models. The dashed lines show the curve without the convex hull. The blue markers on the curves show the operating points where the mentioned cost function is minimized.

The mean and standard deviation of the confusion matrices for the three classification models using random forest models are given in Table 3. The first column shows the loss function that is minimized. Increasing the weight of false negative rate in the loss function, increased the true positive rate (TPR) however it reduced the true negative rate (TNR). For every loss function, the model that used ALLR had better TPR and TNR.

**Table 3.** Confusion matrices of predicting need for ventilation for validation sets of 50 iterations. The values outside the parenthesis show the number of records and the values in parenthesis are the normalized values. The confusion matrices were calculated using thresholds that minimize the cost function mentioned in the first column.

|  |  | Model-CP |  |  | Model-CTSS |  |  | Model-ALLR |  |  |
| --- | --- | --- | --- | --- | --- | --- | --- | --- | --- | --- |
|  |  | Pred -ve | Pred +ve | std | Pred -ve | Pred +ve | std | Pred -ve | Pred +ve | std |
| FPR<br>+<br>0.5*FNR | Actual | 47.66 | 6.34 | 3.94 | 47.34 | 6.66 | 4.60 | 47.74 | 6.26 | 3.54 |
|  | -ve | (0.882) | (0.118) | (0.07) | (0.877) | (0.123) | (0.08) | (0.884) | (0.116) | (0.06) |
|  | Actual | 1.94 | 5.06 | 1.40 | 1.68 | 5.32 | 1.08 | 1.44 | 5.56 | 1.15 |
|  | +ve | (0.28) | (0.72) | (0.2) | (0.24) | (0.76) | (0.15) | (0.21) | (0.79) | (0.16) |
| FPR<br>+<br>FNR | Actual | 43.34 | 10.66 | 5.35 | 42.9 | 11.1 | 5.76 | 44.16 | 9.84 | 5.05 |
|  | -ve | (0.80) | (0.20) | (0.1) | (0.79) | (0.21) | (0.1) | (0.82) | (0.18) | (0.09) |
|  | Actual | 1.18 | 5.82 | 1.07 | 0.9 | 6.1 | 0.96 | 0.74 | 6.26 | 0.87 |
|  | +ve | (0.17) | (0.83) | (0.15) | (0.13) | (0.87) | (0.14) | (0.11) | (0.89) | (0.12) |
| FPR<br>+<br>2*FNR | Actual | 35.66 | 18.34 | 12.87 | 38.94 | 15.06 | 6.52 | 39.36 | 14.64 | 8.81 |
|  | -ve | (0.66) | (0.34) | (0.2) | (0.72) | (0.28) | (0.12) | (0.73) | (0.27) | (0.16) |
|  | Actual | 0.56 | 6.44 | 0.64 | 0.5 | 6.5 | 0.70 | 0.3 | 6.7 | 0.54 |
|  | +ve | (0.08) | (0.92) | (0.09) | (0.07) | (0.93) | (0.1) | (0.04) | (0.96) | (0.07) |

In Supplementary section 4, we provide examples of two patients (one not requiring MV, and the other requiring MV) from the MGMCH dataset demonstrating how the model can be used to forecast outcome.

## Discussion

In this clinico-radiological prediction model for COVID-19 patients from LMIC setting, we showed that synthesis of clinical data with automated CT scan derived lung involvement data (ALLR) performed marginally better in predicting the need for ventilation than similar scores derived from clinical data and traditional CT severity score generated by a radiologist. Importantly, the model output had a low false negative rate, which is important in the context of triage, as a patient with high likelihood of clinical decline should not be triaged as low risk. Therefore, even without a radiologist input, the CT scan could be utilised meaningfully towards model development. This could help in reducing the load on radiologists in generating time-critical reports that incorporate detailed inflammation severity.

The image analysis involves the segmentation of lungs and subsequently diseased areas within the lung tissue. The technique used is robust because they were validated on unseen data, and they were manually examined and the essential postprocessing steps were added to the pipeline in order to make them more accurate. The automated calculation of ALLR from the segmented volumes is a precise method of quantification of abnormality than the CT severity score, independent of observer based variability.

There are multiple features that make this model unique. First, this is the first time that automated CT scan derived data have been amalgamated with clinical features. Second, this takes the clinical and laboratory values on the day of the CT scan and therefore the model represents the clinical picture of the patient on the day of imaging. Third, the outcome of need-for-ventilation is clinically significant, both in terms of clinical outcome and also resource allocation in a healthcare system. Fourth, the underlying segmentation tools were trained using labeled publicly available data from a different setting external to India and the subsequent development of models utilized data from Indian patients. Therefore, the approach and the results are potentially generalizable, at least in other LMIC settings. Fifth, the inclusion of the day-from-symptom-onset potentially adds valuable information about the natural history of the disease (for example, the CT findings on day 7 has a different meaning than the same CT finding on day 21 from pathological and natural-history-of-disease point of view). This is not included in other current clinical models. Finally, the outcome classification results using the ALLR improve slightly over that using the CT severity score. This indicates that the use of the ALLR in place of the CT severity score does not result in any loss in performance. We investigated the possible relationship between the ALLR and the CT severity score. Under linear fit, the correlation coefficient between those was 0.70. A quadratic model provided an improved fit (with root mean squared error reducing to 0.118 from 0.131), and is shown in Supplementary Figure S1.

There are several limitations of the study. First, important physiological parameters on respiratory rate and SpO2 on air presentation were not collected due to the retrospective nature of study. This should be added in the subsequent iterations to make the prediction more robust and most likely will make the model significantly better. Second, Data collected from single centre might create bias in the results that may arise from local clinician practice of selection of patients for mechanical ventilation. Third, The dataset was developed during the first wave of COVID-19 in India. The changing nature and virulence of the virus may alter the performance of the model and recalibration may be required in successive waves. Fourth, the patients were not on therapy at the time of acquisition of the CT scan. Therefore, it is not clear whether the model can be applied to patients already admitted to the hospital and who have been given proven therapy (e.g. systemic corticosteroid or IL-6 inhibitors). Fifth, it should be noted that specific lung pathology cannot be differentiated through this method and therefore a radiologist should still view the images from the standpoint of traditional reporting. Finally, there is a need for a strategy to be devised for scans done in the HRCT format, in order to extrapolate the model developed on volume CT scans into high-resolution scans which are done with fewer sections. Furthermore, there is no detail demographic distribution data regarding the publicly held datasets used in training segmentation models and that could introduce bias, although the effect from demographic and age related variation of lung anatomy in the context of measuring inflammatory burden from COVID-19 is likely to be insignificant.

All these aspects, when taken together, can potentially lead to a robust and standardizable mathematical model to predict individual patient level outcomes in order to achieve efficient triaging in hospitals and critical care units. It is important to acknowledge here that as in other prediction models, different iterations will be needed in different settings and timings of the pandemic to make this approach useful in future. Future prospective studies with expanded data from multiple centers can improve the generalizability of model output, at the same time making it more robust.

## Conclusions

In conclusion, a clinicoradiological model, developed by amalgamation of radiological and clinical parameters, produced in line with the current study design, can predict important clinical outcome of need for invasive mechanical ventilation efficiently and safely. Every setting or region can use this technique to predict the outcome of severe COVID-19 patients effectively.

## Supporting information

TRIPOD checklist

## Data Availability

The data collected for this study are available from the corresponding authors upon reasonable request.

## Author contributions statement

A.S. developed ML models via experimentation, performed validation, wrote the original draft; S.P.J. performed data collection, radiological assessment of CT scans, reviewed the experimental plan and reviewed the manuscript; P.S.D. contributed to the analysis plan and reviewed the manuscript; S.J. and R.S. conceived the study, provided overall supervision, and contributed to editing and in finalizing the manuscript.

## Code availability

The code related to this study are available from the corresponding authors upon reasonable request.

## References

1. https://www.who.int/news/item/26-11-2021-classification-of-omicron-(b.1.1.529)-sars-cov-2-variant-of-concern.

2. Dong, E., Du, H. & Gardner, L. An interactive web-based dashboard to track covid-19 in real time. The Lancet infectious diseases 20, 533–534 (2020).

3. https://coronavirus.jhu.edu/map.html.

4. Emanuel, E. J. et al. Fair allocation of scarce medical resources in the time of covid-19 (2020).

5. https://www.who.int/ethics/publications/ethics-covid-19-resource-allocation.pdf?ua=1.

6. Schwab, P. et al. Real-time prediction of covid-19 related mortality using electronic health records. Nat. communications 12, 1–16 (2021).

7. Knight, S. R. et al. Risk stratification of patients admitted to hospital with covid-19 using the isaric who clinical characterisation protocol: development and validation of the 4c mortality score. bmj 370 (2020).

8. Yang, R. et al. Chest ct severity score: an imaging tool for assessing severe covid-19. Radiol. Cardiothorac. Imaging 2, e200047 (2020).

9. Hadied, M. O. et al. Interobserver and intraobserver variability in the ct assessment of covid-19 based on rsna consensus classification categories. Acad. radiology 27, 1499–1506 (2020).

10. Bellini, D. et al. Diagnostic accuracy and interobserver variability of co-rads in patients with suspected coronavirus disease-2019: a multireader validation study. Eur. radiology 31, 1932–1940 (2021).

11. Shiri, I. et al. Machine learning-based prognostic modeling using clinical data and quantitative radiomic features from chest ct images in covid-19 patients. Comput. biology medicine 132, 104304 (2021).

12. https://data.worldbank.org/indicator/sh.med.phys.zs?locations=inmost_recent_value_desc=true.

13. https://coronacases.org.

14. Jun, M. et al. Covid-19 ct lung and infection segmentation dataset. Zenodo, Apr 20 (2020).

15. Roth, H. et al. Rapid artificial intelligence solutions in a pandemic-the covid-19-20 lung ct lesion segmentation challenge. (2021).

16. An, P. et al. Ct images in covid-19 [data set]. The Cancer Imaging Arch. (2020).

17. Clark, K. et al. The cancer imaging archive (tcia): maintaining and operating a public information repository. J. digital imaging 26, 1045–1057 (2013).

18. Beretta, L. & Santaniello, A. Nearest neighbor imputation algorithms: a critical evaluation. BMC medical informatics decision making 16, 197–208 (2016).

19. Azad, R., Asadi-Aghbolaghi, M., Fathy, M. & Escalera, S. Bi-directional convlstm u-net with densley connected convolutions. In Proceedings of the IEEE/CVF International Conference on Computer Vision Workshops, 0–0 (2019).

20. Caselles, V., Kimmel, R. & Sapiro, G. Geodesic active contours. Int. journal computer vision 22, 61–79 (1997).

21. Marquez-Neila, P., Baumela, L. & Alvarez, L. A morphological approach to curvature-based evolution of curves and surfaces. IEEE Transactions on Pattern Analysis Mach. Intell. 36, 2–17 (2013).

22. Zhou, Z., Siddiquee, M. M. R., Tajbakhsh, N. & Liang, J. Unet++: A nested u-net architecture for medical image segmentation. In Deep learning in medical image analysis and multimodal learning for clinical decision support, 3–11 (Springer, 2018).

23. Breiman, L. Random forests. Mach. learning 45, 5–32 (2001).

24. Chen, T. & Guestrin, C. Xgboost: A scalable tree boosting system. In Proceedings of the 22nd acm sigkdd international conference on knowledge discovery and data mining, 785–794 (2016).

25. Sagi, O. & Rokach, L. Ensemble learning: A survey. Wiley Interdiscip. Rev. Data Min. Knowl. Discov. 8, e1249 (2018).

26. Zhou, Z.-H. Ensemble methods: foundations and algorithms (Chapman and Hall/CRC, 019).

27. Xu, Q.-S., Liang, Y.-Z. & Du, Y.-P. Monte carlo cross-validation for selecting a model and estimating the prediction error in multivariate calibration. J. Chemom. A J. Chemom. Soc. 18, 112–120 (2004).

28. https://www.python.org/.

29. https://colab.research.google.com/.

30. Abadi, M. et al. Tensorflow: A system for large-scale machine learning. In 12th {USENIX} symposium on operating systems design and implementation ({OSDI} 16), 265–283 (2016).

31. https://opencv.org/.

32. Van der Walt, S. et al. scikit-image: image processing in python. PeerJ 2, e453 (2014).

33. Pedregosa, F. et al. Scikit-learn: Machine learning in Python. J. Mach. Learn. Res. 12, 2825–2830 (2011).

