## Supplementary for "An ML prediction model based on clinical parameters and automated CT scan features for COVID-19 patients"

### 1 Data Preparation

The CT scans in dataset-1 and dataset-2 were in NifTI file format. For both of these datasets, axial slices corresponding to each patient scans were saved in PNG image format in individual folders for each patient. The conversion to PNG was required for efficient data loading pipeline for training the models, and the folder structure was created to enable splitting of data into train and test subsets while ensuring that there is no patient overlap between those subsets. To reduce the computational costs, the images were resized to 256x256 using bicubic interpolation.

The CT scans in MGHCH dataset (henceforth called as dataset-3) were in DICOM format. The axial slices were saved in PNG image format, in individual patient folders, after resizing the 3d volume to 256x256x(n/8) using trilinear interpolation where n is the number of axial slices. To make the number of slices a multiple of 8, up to 7 slices of CT scans from the bottom of body were removed. There were two reasons to reduce the z-resolution of images. First, it reduced the computational costs. Second, it increased the effective slice thickness of images and reduced the difference in slice thickness between dataset-2 and dataset-3. All the images were normalized using min-max scaling before feeding to the models.

### 2 Data Augmentation

#### 2.1 Lung Segmentation

The following augmentations were done to the pairs of the input images and labels: random rotations between - 5 to 5 degrees, zoom in to the central 90% of the image 0.5 probability. Gaussian noise was added to the input image with mean 0 and standard deviation 0.1.

#### 2.2 Inflammation Segmentation

The following augmentations were used for input extracted lungs and corresponding inflammation mask while training the UNet++ models: random rotations in the range -10 to 10 degrees, random cropping to central 90% of the image. Gaussian random noise with mean 0 and standard deviation 0.01 was added to the input image.

### 3 Hyperparameters Tuning

The grid search for random forest model was performed over n_estimaters = {100, 200, 300, 400, 500} and max_depth = 1, 2, 3, .., 10, no_limit. The optimal hyperparameters for Model-CP, Model-CTSS, and Model-ALLR were {n_estimaters = 350, max_depth = 3}, {n_estimaters = 350, max_depth = 4}, and {n_estimaters = 400, max_depth = 1} respectively.

The grid search for XGBoost was performed over n_estimaters = {200, 250, 300, ..., 500} and max_depth = {1, 2, 3, .., 10, 15}. The optimal hyperparameters for Model-CP, Model-CTSS, and Model-ALLR were {n_estimaters = 400, max_depth = 3}, {n_estimaters = 350, max_depth = 4}, and {n_estimaters = 400, max_depth = 3} respectively.

### 4 Patient Examples

In Table S1, we provide examples of two patients (Patient 1 not requiring MV and Patient 2 requiring MV as an actual outcome) from the MGMCH dataset, and show how the model can be utilized to generate risk profiles for those patients, in order to aid in clinical decision making in real time. According to the model, the likelihood of need for MV was 0.24 (low) and 0.73 (high) in Patients 1 and 2, respectively, assuming equal weights for FN and FP ($\alpha = 1$). Even when FP is weighed twice compared to FN ($\alpha = 0.5$), the aforementioned likelihoods, 0.17 (low) and 0.63 (high), respectively, remain somewhat similar. In general, a system, depending on its resource availability and acceptability of various risk levels, can calibrate the output of the model accordingly.

### Supplementary Tables

| | Patient 1 | Patient 2 |
|---|---|---|
| Age | 25-30 | 75-80 |
| Sex | Male | Female |
| Day of presentation | 6 | 5 |
| CRP | 4.5 | 165 |
| Ferritin | 85 | 1426 |
| D dimer | 232 | 394 |
| Diabetes | No | No |
| Hypertension | No | Yes |
| Obesity | No | Yes |
| CT severity score | 3 | 24 |
| ALLR | 0.04 | 0.71 |
| Probability ($\alpha = 0.5$) | 0.17 | 0.63 |
| Probability ($\alpha = 1$) | 0.24 | 0.73 |
| Actual need for ventilation | No | Yes |

**Table S1.** The table shows the recorded details of Patients 1 and 2, as well as the estimated ALLR and the probability predicted by model-ALLR (the input to the model being clinical parameters and ALLR).

#### Supplementary Figures

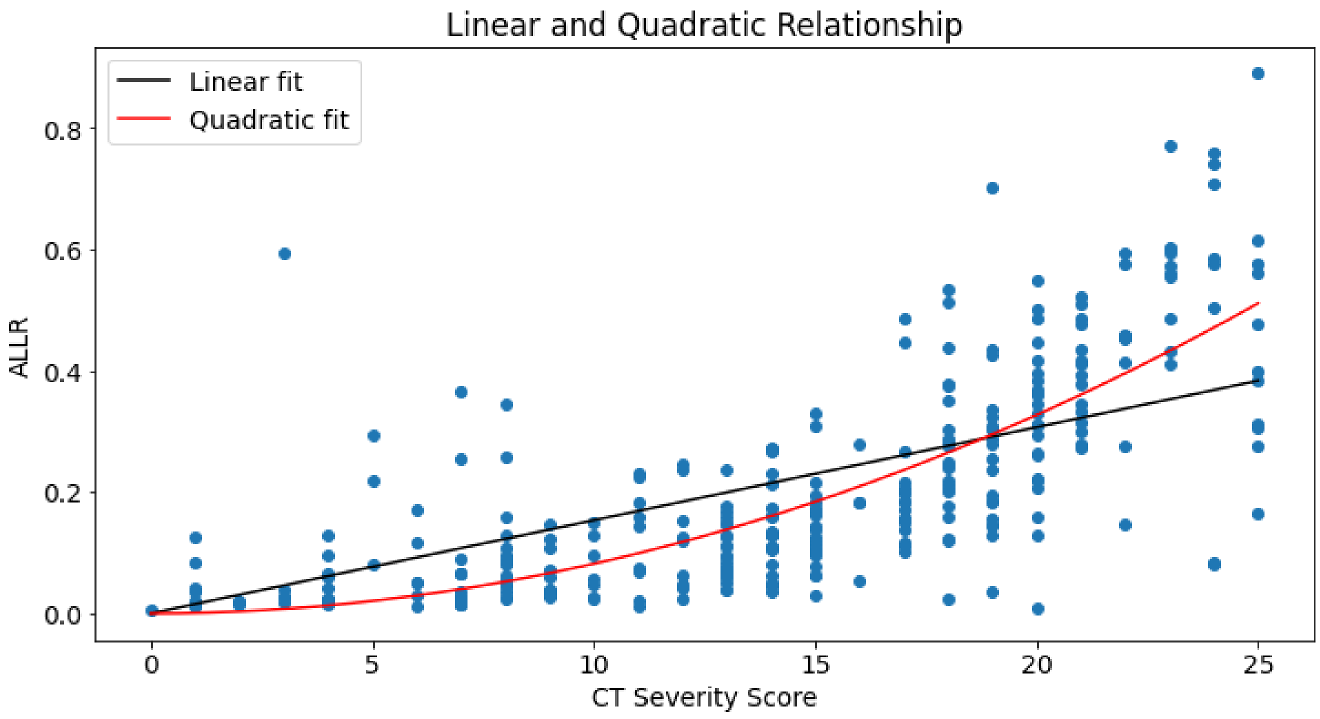


**Figure S1.** Linear and quadratic relationship between CT severity score and ALLR.
